# Guidelines and Standard Frameworks for Artificial Intelligence in Medicine: A Systematic Review

**DOI:** 10.1101/2024.05.27.24307991

**Authors:** Kirubel Biruk Shiferaw, Moritz Roloff, Irina Balaur, Danielle Welter, Dagmar Waltemath, Atinkut Alamirrew Zeleke

## Abstract

A growing volume of evidence marks the potential of Artificial Intelligence (AI) in medicine, in improving diagnostic accuracy, clinical decision support, risk/event prediction, drug discovery, and patient management. However, the continuous integration of AI into clinical settings requires the development of up-to-date and robust guidelines and standard frameworks that consider the evolving challenges of AI implementation in medicine. This review evaluates these guidelines’ quality and summarizes ethical frameworks, best practices, and recommendations.

The Appraisal of Guidelines, Research, and Evaluation (AGREE II) tool was used to assess the quality of guidelines based on six domains: scope and purpose, stakeholder involvement, rigor of development, clarity of presentation, applicability, and editorial independence. The protocol of this review including the eligibility criteria, the search strategy data extraction sheet and methods, was published prior to the actual review with International Registered Report Identifier (IRRID) of (DERR1-10.2196/47105).

The initial search resulted in 4,975 studies from two databases and five studies from manual search. Nine articles were selected for data extraction based on the eligibility criteria. We found that while guidelines generally excel in scope, purpose, and editorial independence, there is significant variability in applicability and the rigour of guideline development. Well-established initiatives such as DECIDE-AI, SPIRIT-AI, and CONSORT-AI have shown high quality, particularly in terms of stakeholder involvement. However, applicability remains a prominent challenge among the guidelines. We conclude that the reproducibility, ethical and environmental aspects of AI in medicine still need attention from both medical and AI communities. This review emphasizes the crucial need for high-quality guidelines and opens a new avenue in evaluating guidelines themselves. Our work highlights the need for working toward the development of integrated and comprehensive reporting guidelines that adhere to the principles of Findability, Accessibility, Interoperability and Reusability (FAIR). This alignment is essential for fostering a cultural shift towards transparency and open science, which are pivotal milestone for sustainable digital health research.

## Introduction

According to the European Union (EU) high level expert group definition, “*Artificial Intelligence (AI) systems are software (and possibly also hardware) systems designed by humans that, given a complex goal, act in the physical or digital dimension by perceiving their environment through data acquisition, reasoning and deciding the best action(s) to take to achieve the given goal*.” [1]. The expert group also described the technical approaches in AI including machine learning (ML) (such as deep and reinforcement learning), machine reasoning (such as knowledge representation) and robotics (such as control, sensors and actuators). In this work, we use the definition of ML and AI proposed by the EU high-level expert group [1].

AI has emerged as a promising and yet disruptive technological advancement with the potential to transform healthcare [2–4]. Studies have shown that AI can improve the diagnostic accuracy, support clinical decisions, predict risk/events, help discover drugs and support patient management [5–7]. Nonetheless, the ongoing incorporation of AI in clinical settings necessitates the development of current, reliable and robust guidelines and standard frameworks that consider the evolving challenges of AI implementation in medicine [8].

Several guidelines for developing and reporting ML models were created by experts worldwide [9, 10]. However, an extensive and continuous evaluation of guidelines is still missing to maintain credibility, standardization, quality of care, patient safety, data protection and ethical research [11]. The agile and ever-evolving challenges in this field impede the process of crafting a gold standard that would cover all aspects of developing and reporting AI studies in the medical domain. For instance, the “hype” in developing and reporting “best performing” models has recently been challenged by questions regarding reproducibility, explainability, governance and, ethical implications for use in healthcare [12]. GenAI (Generative-AI) and LLMs (Large Language Models) have already stimulated substantial discourse in science and innovation since 2022 [13].

Reproducibility is one of the most prominent challenges for AI in medicine, and science in general. Often general textual descriptions of methods and results are published, with over-simplistic levels of details about the necessary steps in pre-processing, model training and validation, and reporting [14]. A limited use of standardized ML model development and reporting guidelines but also the lack of standardised sharing practices of input data and source code hamper reproducibility [15, 16]. From a computational modelling point of view, sharing the model data and code would foster the reusability of the models to answer new research questions or advance the performance of the existing models [16], a topic that has long been discussed in other fields such as Systems Medicine.

Due to its complex and sensitive nature, experts and regulatory stakeholders continuously seek up-to-date guidelines when applying AI in medicine. Often, fragments of suggestions and guiding frameworks are developed by different experts, and scientists face the challenge of choosing the appropriate guideline for a specific use case [17]. Thus, the evaluation of existing guidelines would help scientists to identify the best framework to follow in a specific project. Here, we performed a systematic review of available guidelines for ML model development and reporting. We assessed the quality of the guidelines and summarize the ethical frameworks, checklists, best practices, and recommendations. We strongly suggest that in addition to developing and updating guidelines, well established datasets and code-sharing concepts should be implemented to harness the benefits of AI in medicine. The Findable, Accessible, Interoperable and Reusable (FAIR) guiding principles are the widely accepted approach for scientific data management and stewardship [18]. Their applicability in making software [19] and digital artifacts such as ML models [20] FAIR has been shown over the past years, and it is evident that adherence to these principles maximizes research value and fosters open and reproducible science [21, 22].

## Methods

A systematic review was conducted following the Preferred Reporting Items for Systematic Review and Meta-Analysis (PRISMA 2020) guidelines [23]. PubMed and WOS databases were systematically searched. Two reviewers screened titles, abstracts, and full texts for eligibility and performed data extraction based on a predefined data extraction sheet. Quality assessment was performed using the AGREE II tool [24] and discrepancies were resolved through consensus or third-party arbitration. Data synthesis and analysis were conducted using Python.

### Protocol and registration

The protocol is published in JMIR Protocols with Digital Object Identifier (DOI) and International Registered Report Identifier (IRRID): DERR1-10.2196/47105 [25].

### Eligibility criteria

All available guidelines, standard frameworks, best practices, checklists and recommendations were included irrespective of the study design. Studies were limited to English language and publications until June 2023.

### Search strategy

A systematic literature search was commenced using medical subject headings (MeSH) terms and keywords for medicine, guidelines and ML (S-4 Supplementary file.docx). We used the PubMed and Web of Science databases and the EQUATOR network, which is a global initiative working towards improving research value by promoting robust reporting guidelines (http://www.equator-network.org/). Google Scholar search for references in selected papers led to more thorough search results. Afterward, the search results were uploaded to an online systematic review tool (Rayyan) and then processed with CADIMA [26], a free web tool facilitating the development of systematic reviews and associated documentation, for further screening and preliminary analysis.

### Study selection

After removing duplicates using CADIMA, titles and abstracts were scanned by two independent reviewers (KBS and MR). The reviewers then performed an independent review of full texts and final decisions on whether to include the article for data extraction were made after discussion.

### Data extraction, collection and management

Two independent reviewers (KBS and MR) extracted relevant information from the identified publications using a predefined information extraction sheet, which gathers pertinent information such as study characteristics, study type, aspect, and specific disease/condition of interest if available.

### Quality and risk of bias assessment

Quality, specifically in the context of guidelines, frames the methodological parameters that dictate how other studies should be conducted, reported and communicated. We assessed the quality of identified guidelines using the AGREE II tool [27]. AGREE II measures the quality of guidelines in six fundamental domains including methodological rigorousness and transparency of the guideline development process [24]. Specifically, AGREE II contains 23 items, each rated on a Likert scale rating from 1(Strongly disagree) to 7(-Strongly agree) and grouped within the following six domains:

*Domain 1-* Scope and purpose: assesses whether the guideline stated the main target and scope of the intended use of the guideline.

*Domain 2-* Stakeholder involvement: assesses whether the guideline development process incorporated a representative view of relevant stakeholders including users.

*Domain 3-* Rigour of development: evaluates the methodological thoroughness followed during the guideline development process.

*Domain 4-* Clarity of presentation: assesses the clarity of format and language conveyed in the proposed guideline.

*Domain 5-* Applicability: assesses the presentation of facilitators and barriers to implement the guidelines. The measures need to be considered for the applicability of the guideline.

*Domain 6-* Editorial independence: assesses the statement with respect to funding bias and competing interests.

*Overall assessment:* This domain reflects the subjective assessment of the evaluators regarding the overall quality of the guideline and their opinion in recommending the use.

### Analysis of the guideline quality assessment

To evaluate the risk of bias, four independent appraisers performed a quality evaluation of the nine identified guidelines. The rating was calculated by scaling the total as a percentage of the maximum possible scores for a specific domain [24]. For example, domain one (scope and purpose) has 3 items. Hence the maximum possible score is 7*3*4 = 84 (where 4 is the number of appraisers), and the minimum possible score is 1*3*4 = 12. Thus, a domain score is calculated as:

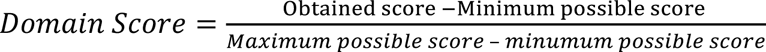

It is important to note that each domain score is calculated independently, and it is neither recommended to combine domains nor to average the result. Item eleven and sixteen, which are specific to medical practice guidelines, were adjusted to the median value for all reviewers. We used Intraclass Correlation Coefficient (ICC) to assess the inter-rater agreement.

## Results

The initial search resulted in 4,975 studies from PubMed and Web of Science (WOS) databases, with additional five studies identified through manual searches in the EQUATOR network and by citation tracking. Two reviewers independently conducted full text reviews of 109 studies and selected nine studies for detailed data extraction and synthesis (Figure 1).

**Figure 1:**
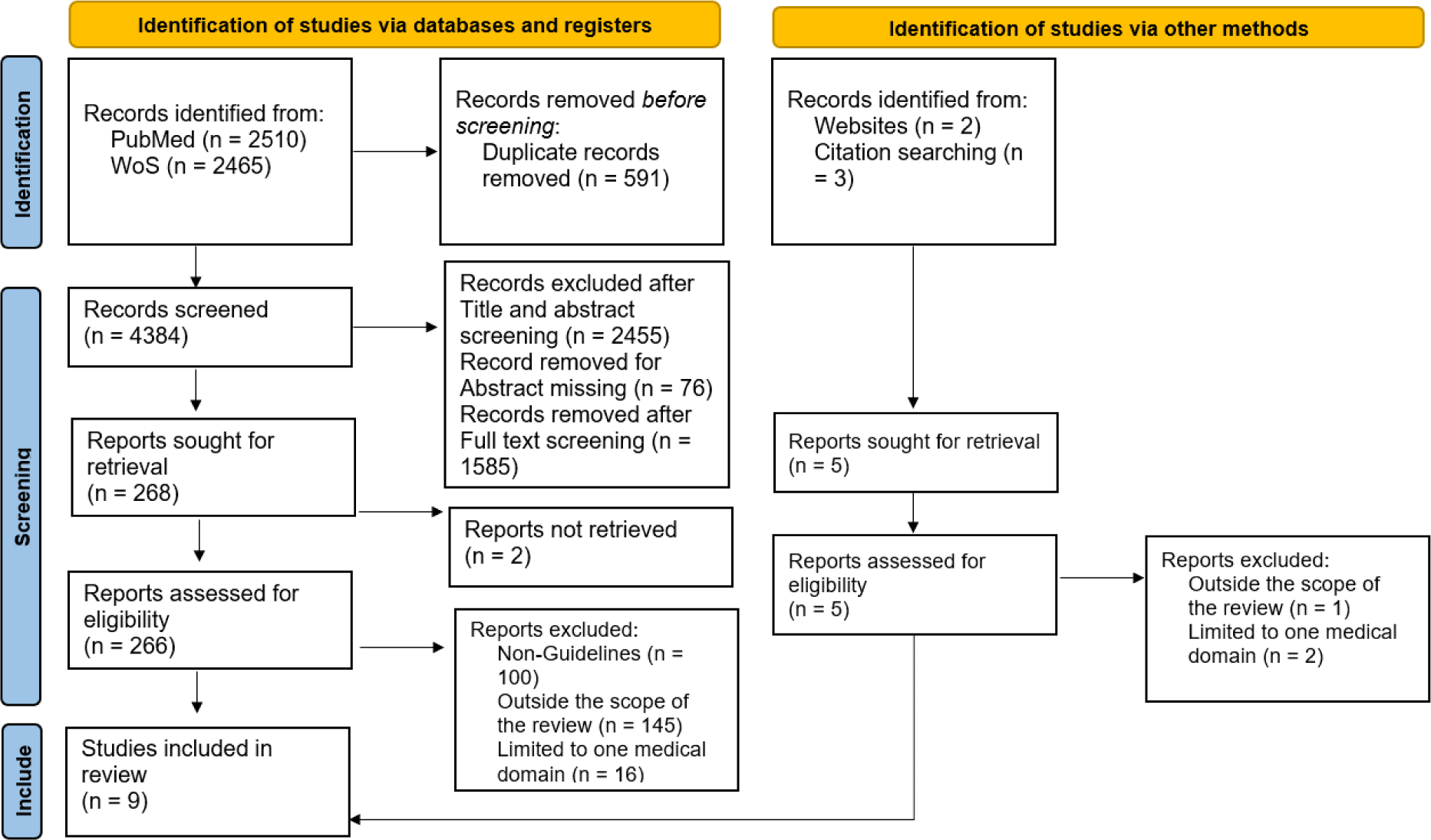
PRISMA Flowchart: Reporting guidelines of AI-related studies in medicine. Reference date: June 2023

Table 1 indicates the main characteristics of the nine reporting guidelines. More details about the selected reporting guidelines are presented in (S-1-Supplementary file.docx) supplementary file.

**Table 1:** Characteristics of the selected reporting guidelines of AI-related studies in medicine.

|  | Guideline | First Author's Name, year of publication | Name of the Journal | Outcome | Aspect | Standard followed | Domain |
| --- | --- | --- | --- | --- | --- | --- | --- |
| 1 | TRIPOD | Gary S Collins, 2015 | Circulation | Checklist of 22 items | Reporting guideline | Yes, Guidance for Developers of Health Research Reporting Guidelines [28] | Diagnostic or prognostic prediction model reporting. TRIPOD AI is under development (The protocol is published, 2021) |
| 2 | SPIRIT-AI | Samantha Cruz Rivera, 2020 | The Lancet Digital Health+ | SPIRIT 2013 items () + 15 checklist items | Trial /Protocol registration | Yes, the EQUATOR Network's methodological framework | Reporting of AI trial protocols |
| 3 | MG | Romana Haneef, 2022 | Archives of Public Health | Checklist of 8 core items with 33 sub items and free text items | Developing and reporting guideline | No, but used a "Step-wise approach" | Reporting linked data/ML studies |
| 4 | CONSORT-AI | Xiaoxuan Liu, 2020 | The Lancet Digital Health+ | CONSORT + 14 new items | Reporting guideline for AI intervention trials | Yes, the EQUATOR Network's methodological framework | Reporting clinical trials with AI intervention |
| 5 | GDRML | Wei Luo, 2016 | Journal of Medical Internet Research | 12 checklist items | Developing and reporting guideline | No/ not reported | Developing and reporting ML studies |
| 6 | STARD 2015 | Patrick M. Bossuyt, 2015 | Radiology+ | 30 checklist items | Reporting guideline | Not reported but used a step wise approach (documented in Equator network) to update the STARD 2003 | Reporting of diagnostic accuracy studies |
| 7 | DECIDE-AI | Baptiste Vasey, 2022 | Nature Medicine | 27 checklist items (17 AI specific and 10 Generic) | Reporting guideline | Yes, the EQUATOR Network's methodological framework | Reporting of early-stage clinical evaluation of AI systems |
| 8 | MI_CLAIM | Beau Norgeo 2020 | Nature Medicine | 21 checklist items | Reporting guideline | Not reported | Reporting best practice checklist for minimum information about AI modeling |
| 9 | CLEAR | Burak Kocak, 2023 | Insights into Imaging | 58 checklist items | Reporting guideline | No/Not reported, but used A modified Delphi method for final selection of items | Reporting guideline for radiomics studies |

+: published in multiple journals

### Quality assessment of the identified guidelines using AGREE II

The Appraisal of Guidelines, Research and Evaluation (AGREE II) tool has been designed to evaluate the quality of clinical practice guidelines [24]. We found that the structured and generic framework can be adapted and applied effectively to evaluate non-clinical practice guidelines [29]. The six core domains of AGREE II are universally applicable to any set of guidelines or recommendations (details in Methods), except for two items (Item 11 and 16), which are specific for clinical practice guidelines. Its standardized evaluation process permits comparable and consistent evaluation across several guideline types. By using AGREE II on this work, we aim to contribute to evaluating the quality, relevance, and impact of non-clinical guidelines, making them a valuable resource for decision-makers and stakeholders.

Our primary domains of focus, in order of relevance, are:

1. The rigour of the guideline development process, (AGREE II Domain 3). We chose it because the thoroughness of the method followed in developing the guideline reflects the quality of the guideline itself.
2. Stakeholder involvement (Domain 2), which indicates whether all relevant stakeholders are involved. The premise is that engaging more stakeholders in the development process of a guideline contributes in its quality and usability.
3. Applicability or instruction how the guideline can be used in practice (Domain 5), which shows the guideline practicality.

Following the guideline assessment, we calculated the aggregated score for each domain by scaling the total (obtained from the 4 reviewers) as a percentage of maximum possible scores (details in Methods).

Our results show that the aggregate scores of Scope and purpose/Domain 1 (which range from 68.1% - 93.3%) and the Editorial independence/Domain 6 (range from 75.0% - 97.9%) are the most satisfied criteria of AGREE II across the guidelines. The aggregated, scaled result of identified guidelines across AGREE II domains is shown in Figure 2: from high percentage value (green) to low percentage value (red).

**Figure 2:**
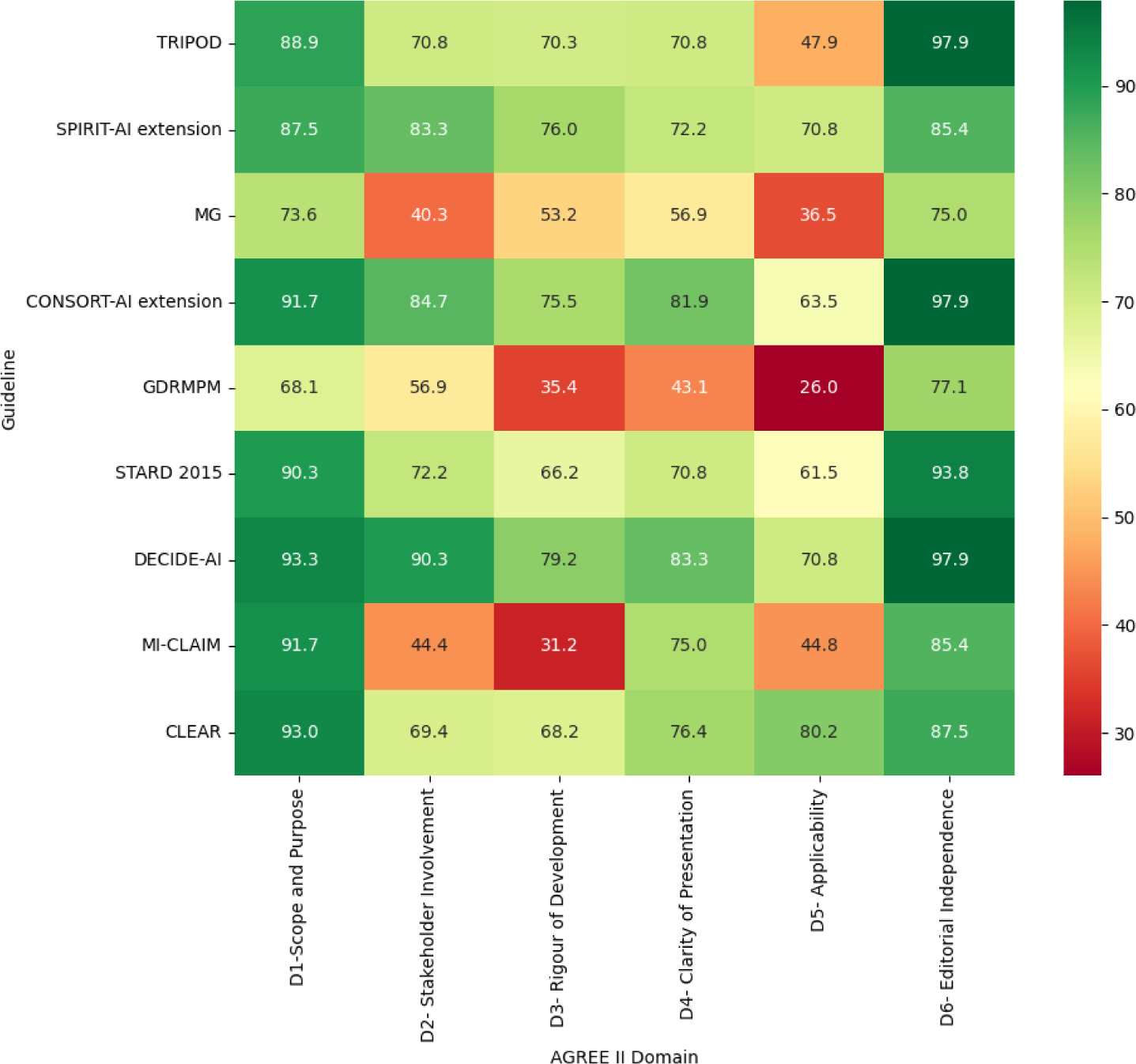
**Heatmap of aggregate scale of AGREE II scores among guidelines with respect to domains.**

We also observed a clear variability of domain scores across guidelines (Figure 3); the lowest scoring domain refers to Domain five/Applicability with 26.0%. When comparing at guideline-level, DECIDE-AI has the highest score across most of the domains. Regarding the “Rigour of development” (Domain 3), notably, only four guidelines scored above 70% (DECIDE AI, CONSORT AI, SPIRIT AI, and TRIPOD), indicating their higher quality.

**Figure 3:**
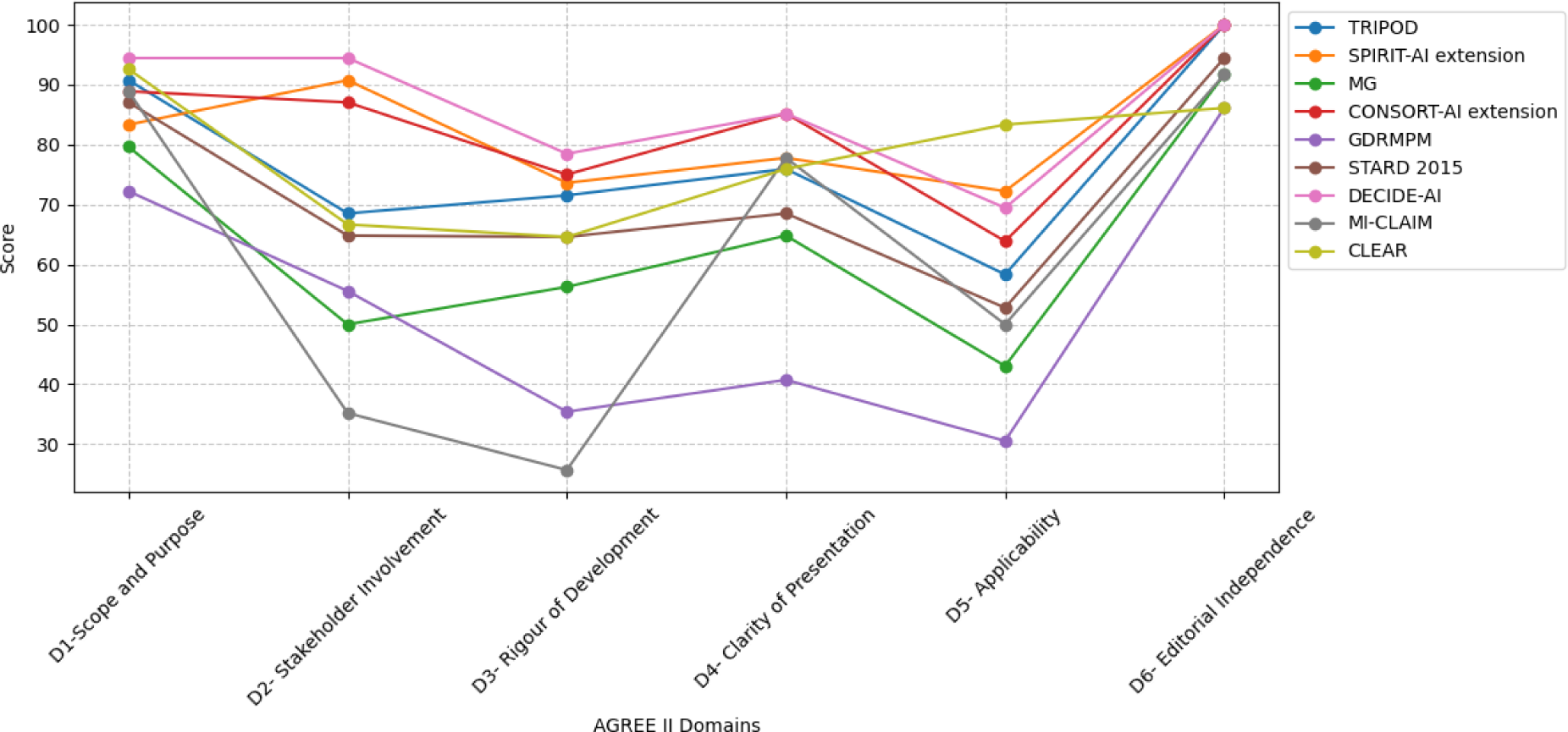
**Parallel coordinate plot of AGREE II domains and guidelines**

DECIDE AI is the highest quality guideline with respect to rigour of development and stakeholders’ involvement, followed by SPIRIT AI and CONSORT AI. Moreover, the guideline CLEAR was scored as the highest quality with respect to applicability. See Figure 4 for more details.

**Figure 4:**
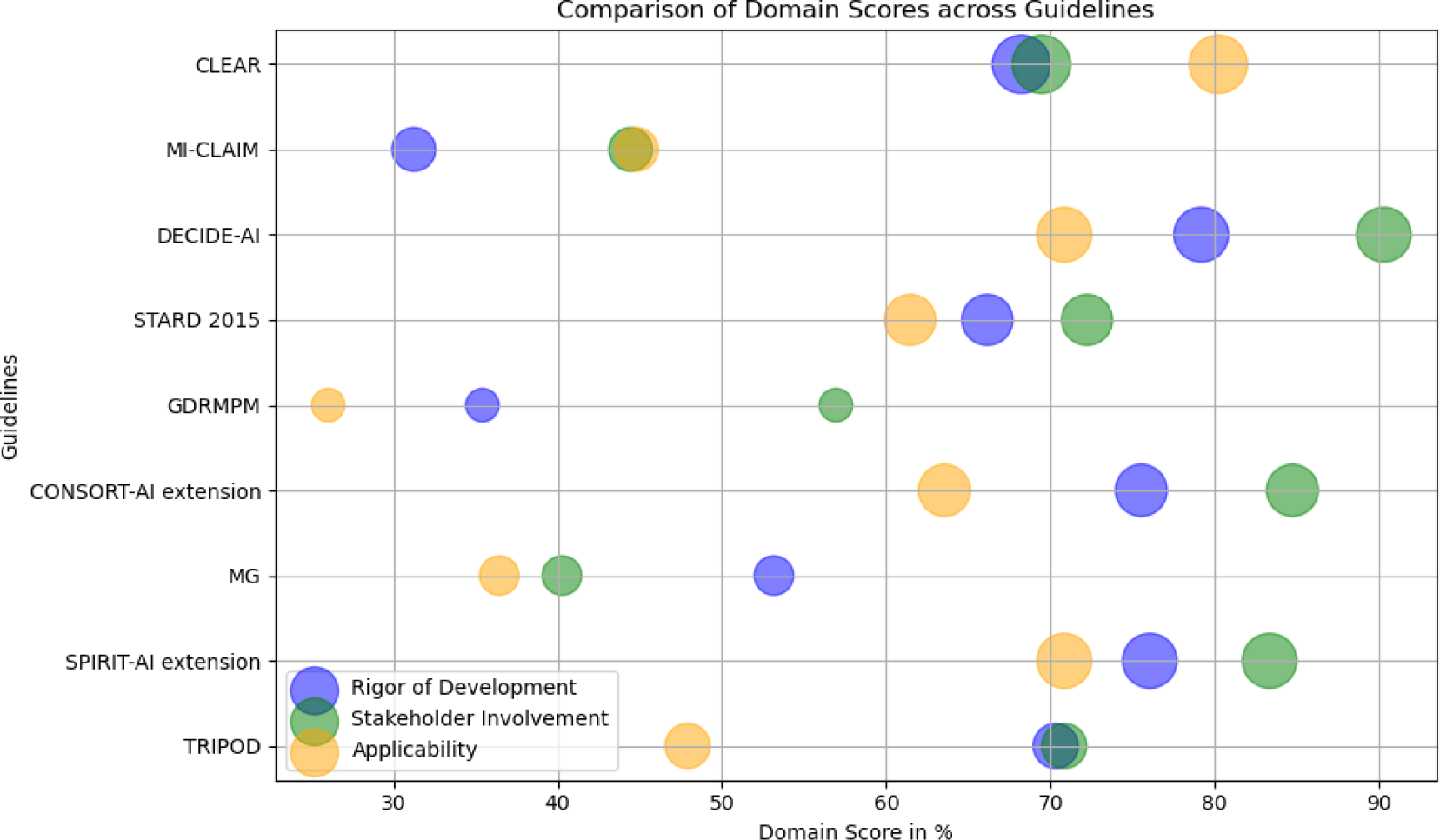
**Scatter plot of targeted AGREE II domains across guidelines.**

We evaluated the inter-rater agreement regarding the overall and primary domain level consensus among the four evaluators using the Intra-Class Correlation Coefficient (ICC) [30]. The overall agreement among the four independent evaluators regarding the guidelines’ quality were statistically significant ranging from ICC of 0.62 – 0.92 with p-value<0.05. The details of individual scoring and domain level ICC can be found in (S-2-Supplementary file.docx) supplementary file.

## Discussion

The systematic search resulted in nine reporting guidelines for AI in medicine. The quality assessment result indicated that the overall quality of available reporting guidelines with respect to describing the scope and purpose (Domain 1) and editorial independence is relatively well scored across the guidelines. Greater variability of scores in explaining the applicability (Domain 5) and rigour of the guideline development process (Domain 3) were observed. With respect to the primary domain of quality evaluation in this study (Domain 3), DECIDE AI, SPIRIT AI and CONSORT AI reporting guidelines scored the highest with 79.2%, 76% and 75.5%, respectively. The secondary quality criterion, stakeholder involvement (Domain 2) was also scored higher by the same guidelines, with score of 90.3%, 83.3% and 84.7%, respectively.

All of the identified guidelines present a way of reporting studies as a checklist of important sections such as introduction, methods, results, discussion, conclusion and additional information sections. The majority of the reporting guidelines were not designed for AI studies *per se* but were extended to accommodate studies involving AI. The extension was mostly done by adding additional items to the checklists that were already in use for reporting a certain type of research findings. For instance, both SPIRIT AI and CONSORT AI are extensions of SPIRIT and CONSORT statements which were originally designed to report clinical trial protocols and clinical trial studies respectively [31].

The “Rigour of development” feature assesses whether the following components are clearly stated in the guidelines: a systematic evaluation of evidence synthesis, method of developing the guideline, explicit link between the guideline and the body of evidence, external expert revision of the developed guideline and the procedure to update or modify the guideline is clearly stated in the suggested guidelines [24]. Most of the identified guidelines have not considered a systematic synthesis of previous works. All guidelines have a justified rationale of their purpose and scope, whereas only half of them followed a standardized procedure of guideline development process such as the one suggested by the EQUATOR Network. However, not following a standard procedures for developing guidelines could result in compromised quality of guidelines [32].

A recent publication [10] reviewed the contents of AI guidelines using translational stage of surveillance domains. The authors showed that most guidelines discussed the importance of ethics, reproducibility and transparency in AI studies but were less likely to engage relevant stakeholders such as patients, end users and experts during the development process. This result is in line with our findings. To engage relevant stakeholders in the process of developing guidelines helps in converging efforts and maximize the utility and versatility of the guidelines [33]. Specifically, DECIDE AI, CONSORT AI and SPIRIT AI guidelines involved a wide range of stakeholders during their development process, while other guidelines were developed by experts and researchers from different institutions without the engagement of potential stakeholders.

Applicability is another important gap in the identified guidelines. To ensure a guideline’s applicability, it is essential to provide a comprehensive description of the factors that facilitate or hinder its application. This can be a detailed presentation of suggested tools and instructions for using the guidelines effectively. It is important to outline the resource implications of applying the guidelines. Furthermore, monitoring or auditing criteria should be explicitly presented to ensure the quality and adherence of a guideline [24]. CLEAR, which is the most recent reporting guideline for radiomics research, [34] was one of the most applicable reporting guidelines in our review. The issue of applicability is not limited to guidelines’ quality but also limited to study design. For instance, most of the quality guidelines that are widely in use in reporting AI related studies are focused on clinical trials or specific fields of study. In contrast, most of the studies in medical contexts applying AI methods are observational studies, which consequently creates a reporting gap in these types of studies. Thus, we strongly suggest that a reporting guideline for AI studies in medicine, irrespective of the study design, should be developed to enhance transparent reporting, reproducibility and reusability, which ultimately contributes to improved healthcare.

Other important aspects of AI applications in medicine are the moral dimensions such as bias, ethics and governance, which are still prominent challenges strongly influencing the deployment of AI systems. A solution proposed by researchers is to embed AI ethics in the entire AI model development process [35].

One aspect of AI that is usually overlooked is its environmental implications [36]. According to the characterizations of the carbon footprint of AI computing considering the lifecycle across large-scale use-cases, the carbon emission to train a ML model is considerably high [37, 38]. We suggest that future reporting guidelines should include a checklist that encompasses the moral and environmental aspects of AI studies as well.

### Contributions to better reproducibility in AI in medicine and beyond

Reproducibility, described as “the ability of an independent research team to produce the same results using the AI method based on the documentation made by the original research team” [15], requires an exact representation of all relevant aspects of the study development and realization. This includes the complete information of the used software and source code, the original data as well as the correct documentation of crucial details and precise instructions for the implementation [39–41]. The reproducibility in AI builds trust in the developed models and results [14, 40]. Therefore, aiming for reproducibility, focusing on the correct and detailed documentation, and providing the necessary details regarding the source code and data information should be mandatory for every researcher and developer to achieve highly valued and trustful scientific findings.

Given its definition, model reproducibility comes with its challenges related to access to data, code, documentation, and clear instructions. Without the opportunity to access any of the given requirements, researchers fail to reproduce roughly similar findings compared to the original study. The lack of proper upkeep of essential resources, such as data, code, or instructions, hinders advancements in research and impedes reproducibility [42]. In addition, the current academic environment encourages researchers to publish prototypes of their AI models rather than ensuring a fully verified system [41], which also impacts the quality of these models. Figure 5 illustrates the three important elements of medical AI research.

**Figure 5:**
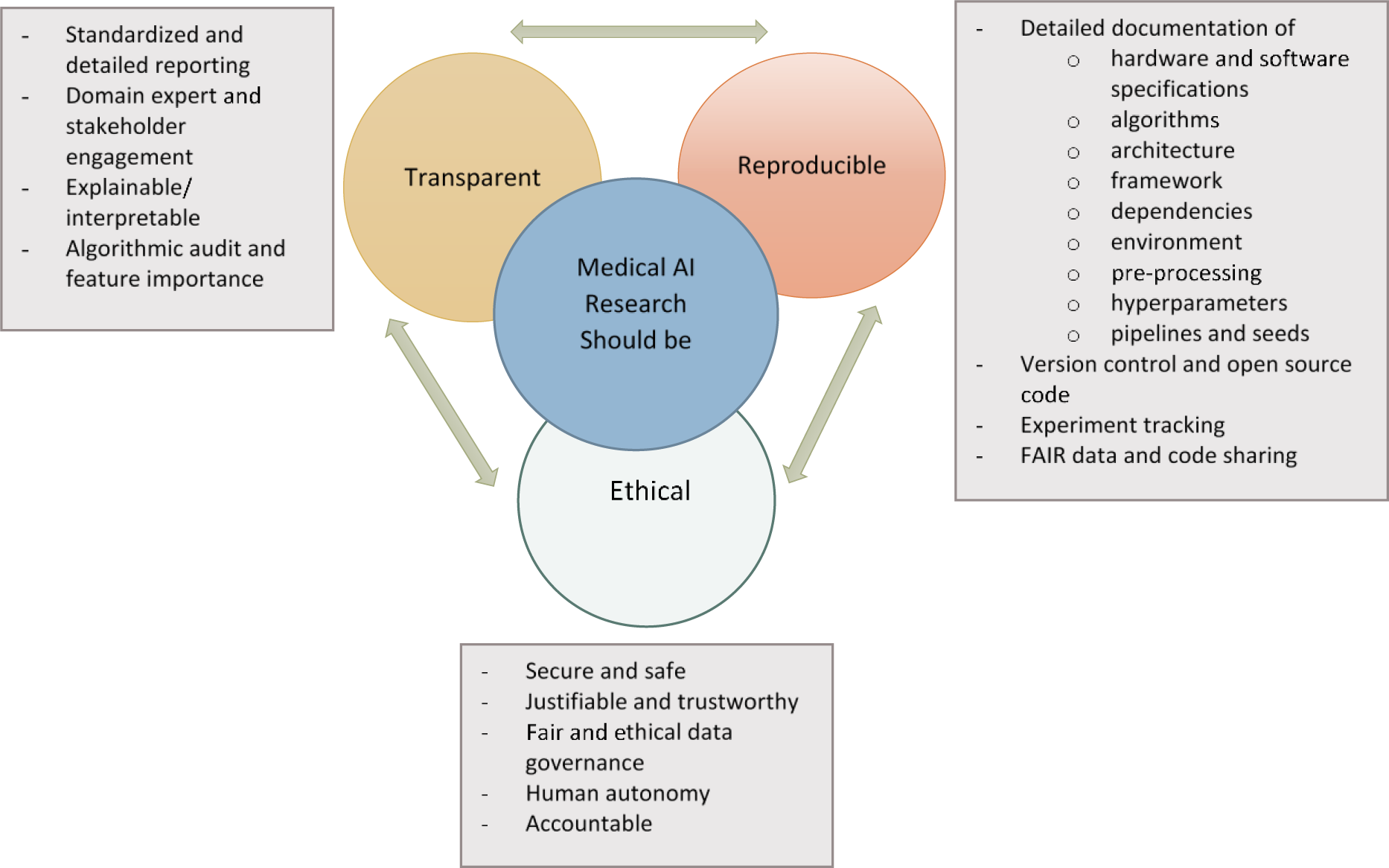
Identified elements of transparency, reproducibility and ethics in medical AI research.

### Standard frameworks and best practices for AI model reproducibility

Frameworks, guidelines, and best practices should offer guidance to achieve a minimum reproducibility standard to ensure reliable results in future studies [43].

Heil et al 2021 [40] proposed a reproducibility standard at three different levels. According to this work, the level of reproducibility can be given on a time-scale based on the time needed to reproduce the work. The scale starts at “forever” for an irreproducible study and ends at “zero” for an automated and fast reproducible study. On this scale, the three degrees “bronze”, “silver”, and “gold” define which requirements have to be met to achieve reproducibility, with “bronze” symbolizing the bare minimum and “gold” meaning the research team ensured full automation. The checklist for reproducibility focuses on a detailed description (and publication) of all used models and algorithms and the complexity of the analysis [44]. Furthermore, any theoretical claim has to be proven entirely and assumptions explained. Figures and tables, as well as the corresponding datasets, have to be described and explained in full, and the work flow should be presented in detail.

### Conclusions

Reporting guidelines are essential when publishing or communicating research results. However, the quality of reporting guidelines should also be evaluated to facilitate harmonization and standardization in communicating research findings [29]. Therefore, our study has assessed reporting guidelines for AI studies in medicine, highlighting their strengths and weaknesses.

Currently, there is no quality assessment tool for measuring the quality of non-clinical practice guidelines involving AI. Therefore, we believe that the development of a quality assessment measure for non-clinical practice guidelines including reporting guidelines should be considered to improve the quality of future guidelines.

The adaptation of guidelines for AI studies clearly improve the completeness of the report. One step towards reproducibility is the publication of code and related information (see the list of resources for sharing code in supplementary file (S-3-Supplementary file.docx).

While these guidelines provide a road map to reproducibility, they also highlight the need for a cultural transformation within the medical AI research community. This change should prioritize transparency, quality, and exhaustive documentation over the rush to publish findings. Additionally, journals should take more responsibility and enforce reproducibility for future AI studies [14]. By doing so, they support efforts to establish a standard within the framework of reproducibility and promote sustainable and transparent research.

The journey towards complete reproducibility in AI research may be lengthy and complex, but it is a worthwhile endeavor. The rewards are not only for individual researchers but for the entire scientific community and society as a whole, as it assures the dependability and trustworthiness of AI systems, which are increasingly pervasive in our daily lives.

Using AGREE II for non-clinical guidelines has its own limitations. Since it is designed for clinical studies, some of the items in the evaluation metrics may not align perfectly with the non-clinical context. Another limitation is that our review is limited to English language publications and this could result in missing important guidelines developed in other languages.

Applicability remains a challenge, so addressing this gap and developing comprehensive guidelines for various AI study types is essential. Our study pinpoints the critical need for quality guidelines and highlight the potential for FAIR integrated and comprehensive reporting guidelines. A cultural shift towards transparency and journals enforcing reproducibility is vital. Although this journey might seem complex, it ensures the reliability of AI systems, benefiting science and society in general.

## Data Availability

All data produced in the present study are available upon reasonable request to the authors

## Availability of data and materials

All data generated or analysed during this study are included in this article. The python code for generating the plots can be found here (https://github.com/kirubel-Biruk-Shiferaw/Guidelines-and-Standard-Frameworks-for-Artificial-Intelligence-in-Medicine-A-Systematic-Review)

## Acknowledgements

KBS would like to thank the DAAD (German Academic Exchange) for supporting the doctoral research study expenses. AZ acknowledge NFDI4Health.

